# Literature study on the return on investment concerning the implementation of a computerized clinical decision support in a hospital information system

**DOI:** 10.1101/2020.10.30.20223362

**Authors:** Gérard Christian Kuotu, Bride Barth Moukam Lower

## Abstract

**Background:** The objective of this study was to carry out a bibliographic study on the return on investment concerning the implementation of a computerized clinical decision support in a hospital information system.

**Methods:** A bibliographic search was carried out using the PubMed and Google Scholar bibliographic databases. The articles obtained were selected by combining the elements: outlook, net benefits, comparability, types of costs, discount rate, sensitivity analyzes, and measurement of effectiveness. This enabled us to review 498 articles published during our study period, of which 56 were selected.

**Results:** The most commonly available tools are return on investment estimation methods. The data in the literature on the return on investment concerning the implementation of clinical decision support, although documented, are quite mixed.

**Conclusion:** Evaluations such as the econometric approach can be considered to determine if these investments are justified.

## Background

Nobel laureate in economics Robert Solow in a 1987 New York Times interview said, “You can see the computer age everywhere but not in productivity statistics”. As a result, it has become essential to accurately assess the added value of investments in information in general and the health sector. Indeed, clinical decision making is defined as a process of considering and comparing the possibilities, risks, uncertainties, and options to choose a course of action [1,2]. Several studies report that “clinical decision support” improves practitioner performance, providing reminders, summaries, and facilitates coordination of care [3,4,5,6]. They improve the quality and reduce the costs of undue health care [7,8] and others, note a reduction in the length of stay of patients in whom an assisted prescription has been used [9]. But even if most of the players agree on their importance, they nevertheless represent substantial budget lines which have not yet indisputably proven their profitability, which still provokes many debates when it comes to justifying expenditure. corresponding [10]. In a context of scarcity of resources, we are entitled to wonder about the potential return on investment, which could constitute the implementation of a computerized clinical decision support in an information system hospital. Financial benefits generally fall into one of these categories: lower costs, improved productivity (which translates into increased income) and improved competitiveness (which translates into income generation). To account for them we commonly use the return on investment (ROI) which is the ratio of the amount of money gained or lost during an investment over the amount of money invested [11]. A positive ROI suggests a quotient of the total benefits provided by a project over the sum of the investments made to achieve it greater than 1. The object of this present study was to carry out a bibliographic study on the return on investment concerning the implementation. implementation of computerized clinical decision support in a hospital information system.

## Material and methods

Between February 28, 2019 and March 30, 2019, we conducted a targeted review of the literature on the return on investment concerning the implementation of a computerized clinical decision support in a hospital information system using the Joanna Briggs Institute Targeted Reviews Guide [Peters et al. (2015)]. Our search was limited to the PubMed^1^ and “Google schoolar”^2^ electronic databases. As a first step, we did not include the term “return on investment” as a search term to allow a broad search, which would include all published articles about decision aids. This search strategy included the terms of following searches: “decision support systems, clinical”, “clinical decision support systems”, “decision support systems, clinical”, “decision support, clinical”, “clinical decision supports”, and allowed us to identify the key words MeSH that we have thus combined: Decision support systems, clinical “; “Cost-Benefit analysis”, “decision making, computer-assisted”. Our articles were then selected based on the combination of the following: outlook, net benefits, comparability, types of costs, discount rate, sensitivity analyzes, and measure of effectiveness. This enabled us to review 498 articles published during our study period, of which 56 were selected.

## Result

### Narrative description of the research selection process accompanied by the research selection flowchart

The flowchart below is inspired by the “PRISMA diagram” (12) and presents the selection process, indicating the results of our research, (see Figure 1).

**Figure 1:**
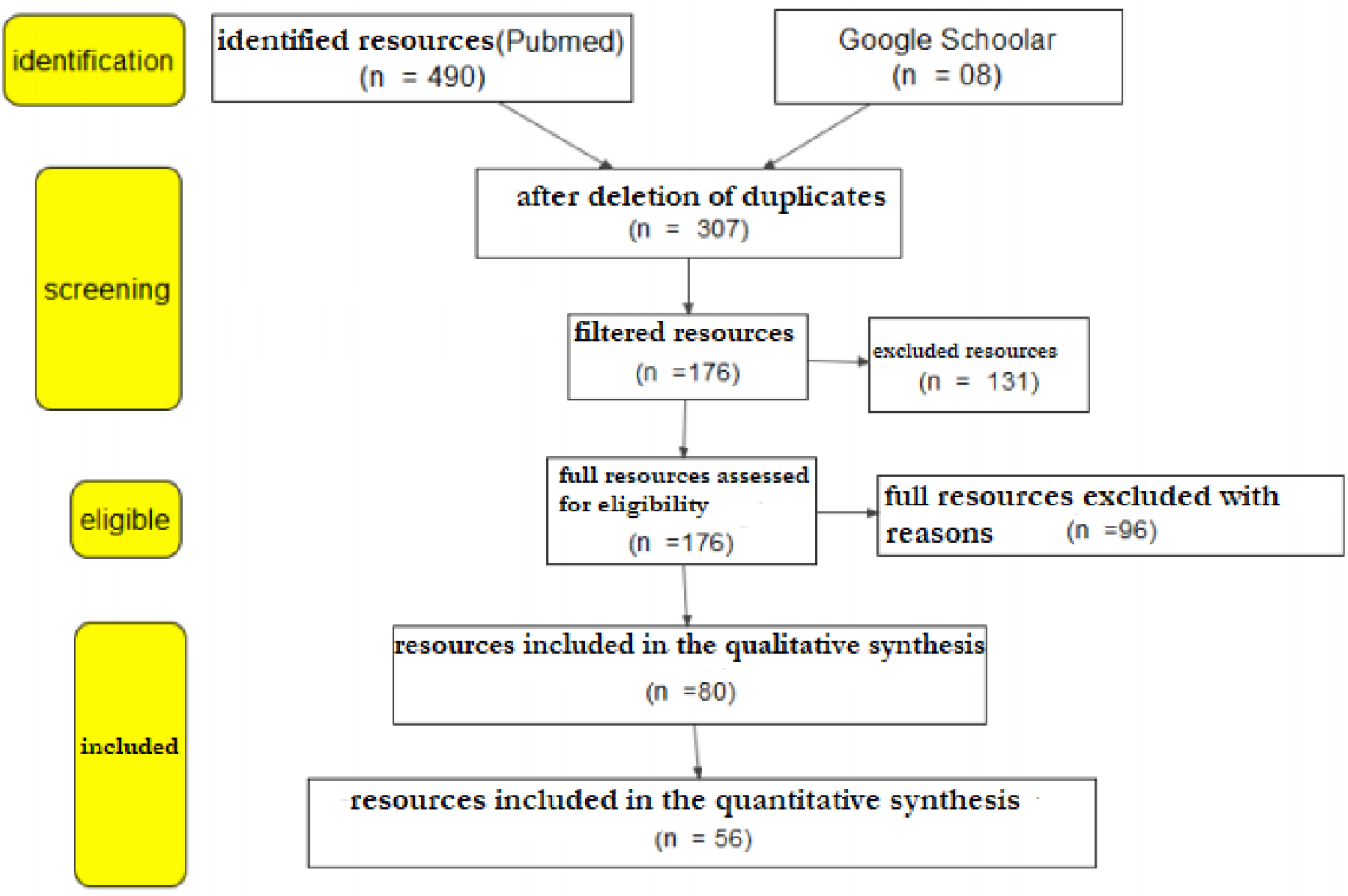
Selection process for references / abstracts of articles. For the bibliographic study on the return on investment concerning the implementation of a computerized clinical decision support in a hospital information system

## Main concepts

### A. Return on investment (ROI) [13.14]

It comes from the financial world and is presented as the ratio between the income generated by an investment and the capital placed in this investment. It is usually expressed as a rate. Thus, if an investment costs 10,000 euros, but brings in 15,000 euros, the ROI will be 50%. It is made up of four main indicators:

1. Net value (NV) and net present value (NPV): which measure, by year and cumulatively, the difference between the consumption of resources and the expected recoverable gains, over the life of the project. The advantage of NPV over NV is that it considers in the calculation the effect of time on value, via the interest rate and monetary erosion or discount rate.
2. The payback period: this is the number of years and months required for the cumulative cash flow to reach the invested capital. It therefore makes it possible to identify from when the project is profitable.
3. The internal rate of return (IRR): this is the rate at which there is an equivalence between the invested capital and the total cash flow. For a project to be acceptable, its IRR must be greater than the minimum rate of return required by the establishment. This rate is called the “rejection rate”. An investment projects is more interesting the higher its IRR.

As we can see, the ROI is a very practical and easy ratio to establish when it comes to studying financially valuable investments.

### B. Determination of qualitative gains (Balanced Scorecard theory) [15]

It is a method developed by “Norton and Kaplan” in the 1990s. Initially focused on private enterprise, it starts from the observation that traditional performance measurement systems, based on only financial indicators, are detrimental to capacity. companies to create long-term economic value. Conversely, the proposed method aims to reflect the value of the company’s intangible assets, its qualitative value, which cannot be financially valued. Norton and Kaplan therefore propose a measurement system based on 4 items:

1. Finance,
2. Performance toward customers,
3. Internal processes,
4. Organizational learning.

It seems particularly well suited to information systems projects hospital, insofar as the 4 items mentioned above are found according to the 4 following axes in a hospital:

1. Improved financial results (cost control, increased revenue),
2. Improvement of the quality and safety of care (patient satisfaction),
3. Improvement of organizations and cooperation between professionals,
4. Improvement of working conditions (satisfaction of hospital professionals).

### Main methodologies [16]

- The EIFIC-HOC tool Developed by the MAINH (National Hospital Investment Support Mission) in the context of the relaunch of the investment required by the 2007 Hospital plan in France, its purpose is to calculate the a priori ROI of a project for several different scenarios and hypotheses.
- Method developed by GMSIH This is a guide developed in 2008 by the “Groupement de modernization des Systèmes d’Information Hospitaliers (GMSIH)”, on the basis of the results of three pilot experiments (at the University Hospital of Amiens, Clermont-Ferrand University Hospital and Rennes in France) to set up a hospital information system. This approach does not only aim to determine the return on financial investment, but also to realistically assess the expected performance of the projects to be carried out in a logic of general optimization.
- The econometric approach Unlike the previous two, the measure of capital efficiency (RSI), is done using cost or production functions.

### Economic analysis

Four elements structure our analysis:

### The definition of the framework of the evaluation

it allowed us on the one hand, to identify which perspective was adopted in the various studies carried out (the company, the patient, health insurance, supplementary insurance, as well as the care establishment …) and secondly, to identify the nature of any comparison made (profitability of the old system compared to the new system …

### Identification, measurement and valuation of costs

three (03) types of costs have been identified: direct medical and non-medical costs (costs of valued strategies taking into account staff, consumables, depreciation of the equipment and its maintenance.), indirect costs (indirect costs for the patient) and intangible costs (cost of anxiety).

### Description of the type of comparative analysis used

this description highlights cost minimization studies, cost-effectiveness studies (years of life saved, etc.), cost-utility studies (quality, etc.), cost studies -Benefit (avoided costs, willingness to pay, etc.).

### Consideration of time and uncertainty

it considers discounting costs and discounting benefits. The summary of the results of this economic analysis are presented in (Table 1):

**Table 1:** Main results of the economic analysis of the return on investment

| **Author,**<br>**Year** | **Evaluation**<br>**framework** | **Type of**<br>**assessment** | **Goal** | **Costs retained** | **Considering**<br>**time** | **Conclusion** |
| --- | --- | --- | --- | --- | --- | --- |
| **Wu RC and**<br>**al.,**<br>**2007 [17]** | Care facility | Cost-effectiveness<br>analysis | To determine the<br>potential return on<br>investment of an<br>aid system<br>electronic | Direct medical<br>and non-medical<br>costs | Yes | RSI >1 |
| **Karnon J and**<br>**al.,**<br>**2008 [18]** | Patients<br>+<br>Care facility | Cost-utility<br>analysis | To estimate the<br>net benefits of<br>interventions<br>aimed at reducing<br>the impact of<br>medical errors | Intangible costs<br>and Indirect costs | Yes | RSI >1 |
| **Rosser WW**<br>**and al.,**<br>**1992 [19]** | Care facility | Cost-effectiveness<br>analysis | To assess the<br>cost-effectiveness<br>of three<br>computerized<br>reminder systems<br>on compliance<br>with tetanus<br>vaccination | Direct costs | Yes | RSI ±1 |
| **Fretheim A**<br>**and al.,**<br>**2006 [20]** | Care facility | Cost-effectiveness<br>analysis | To compare the<br>costs and effects<br>of multiple<br>antihypertensive<br>prescriptions<br>using the standard<br>method, compared<br>to assisted<br>prescriptions | Direct costs | Yes | RSI >1 |
| **Plaza V and**<br>**al.,**<br>**2005 [21]** | Care facility | Cost benefit<br>analysis | To evaluate the<br>cost-effectiveness<br>of a DRMS to<br>promote the<br>SADM<br>recommendation in relation to<br>current practice | Indirect and direct<br>costs | Yes | RSI±1 |
| **Chisolm DJ**<br>**and al.,**<br>**2006 [22]** | Patients<br>+<br>Care facility | Cost benefit<br>analysis | To evaluate the effectiveness between a computerized order and a standard order in the pediatric asthma care process | Intangible costs | Yes | RSI±1 |
| **Mekhjian**<br>**HS and al.,**<br>**2002 [23]** | Patient Health<br>Insurance | Cost benefit<br>analysis | To evaluate the advantages of an expert system on health care delivery | Indirect and<br>intangible costs | Yes | RSI±1 |
| **Stone WM**<br>**and al.,**<br>**2009 [24]** | Patients | Cost benefit<br>analysis | To evaluate the implementation of an Expert system in the care of operated patients | Indirect costs | Yes | RSI >1 |
| **Tierney**<br>**WM.,**<br>**1993 [25]** | Care facility | Cost benefit<br>analysis | To support IT costs and reduce costs | Indirect and<br>intangible costs | Yes | RSI±1 |

## Discussion

The aim of our work was to carry out a bibliographic study on the return on investment concerning the implementation of a computerized clinical decision support in a hospital information system. It emerges that to perform this analysis, the most commonly available tools are methods of estimating the return on investment [26]. Several interesting studies have shown the value of these analyzes in hospitals. However, by their intrinsic nature (financial / economic) the previous methods do not consider the less tangible aspects of the impact of these decision aids. As alternatives, recent studies have shown the value of the econometric approach in evaluating the impact of hospital information systems [27,28,29,30]. However, our study has several limitations. First, the divergent assessment methodologies from one country to another, from one hospital to another do not allow us to come up with a single methodology. As a result, most health establishments cannot carry out a study given the necessary resources, which are significant.

## Conclusion

The data in the literature on return on investment concerning the implementation of clinical decision support, although documented, are quite mixed. Some studies have shown that these “expert systems” could offer cost advantages despite ever higher acquisition costs, while others find no added value to justify their use. However, given the uncertainty surrounding the data on the costs and potential gains around their implementation, it is difficult to draw a definitive conclusion as to whether the current return on investment may justify investing more in these. technologies. Evaluations such as the econometric approach in evaluating the impact of hospital information systems can be conducted to determine whether these investments are justified.

## Data Availability

not applicable

## Author’s contribution

All authors contributed to the study conception and design. Gerard Christian KUOTU, had the idea for the article and performed the literature search, Gerard Christian KUOTU and performed the analysis, Gerard Christian KUOTU, and Bride Barth MOUKAM LOWER drafted the article, and Gerard Christian KUOTU critically revised the work.

## Compliance with ethical standards Conflict of interesse

None.

## Ethical standards

We confirm that the approval of an institutional review board was not required for this work and we have read the Journal’s position on issues involved in ethical publication and affirm that this work is consistent with those guidelines.

## Additional material’s

## Footnotes

1 https://www.ncbi.nlm.nih.gov/pubmed

2 https://scholar.google.com/

## References

[1] Croskerry P. The importance of cognitive errors in diagnosis and strategies to minimize them. Acad Med 2003;78(8):775–80.

[2] Elstein AS, Schwarz A. Clinical problem solving and diagnostic decision making: selective review of the cognitive literature. BMJ 2002;324(7339):729–32.

[3] Anderson JA, Willson P, Peterson NJ, Murphy C, Kent TA. Prototype to practice: developing and testing a clinical decision support system for secondary stroke prevention in a veterans healthcare facility. Comput Inform Nurs. 2010;28:353–363.

[4] Montgomery AA, Fahey T, Peters TJ, MacIntosh C, Sharp DJ. Evaluation of computer based clinical decision support system and risk chart for management of hypertension in primary care: randomised controlled trial. BMJ. 2000;320:686–690. 9

[5] Garg AX, Adhikari NK, McDonald H, Rosas-Arellano MP, Devereaux PJ, Beyene J, Sam J, Haynes RB. Effects of computerized clinical decision support systems on practitioner performance and patient outcomes: a systematic review. JAMA. 2005;293:1223–1238.

[6] Shojania KG, Jennings A, Mayhew A, Ramsay CR, Eccles MP, Grimshaw J. The effects of on-screen, point of care computer reminders on processes and outcomes of care. Cochrane Database Syst Rev. 2009 Jul 8;3:CD001096.

[7] Bosworth HB, Olsen MK, McCant F, Harrelson M, Gentry P, Rose C, Goldstein MK, Hoffman BB, Powers B, Oddone EZ. Hypertension Intervention Nurse Telemedicine Study (HINTS): testing a multifactorial tailored behavioral/ educational and a medication management intervention for blood pressure control. Am Heart J. 2007;153:918–924.

[8] Barnett GO, Cimino JJ, Hupp JA, Hoffer EP. DXplain - an evolving diagnostic decision –support system. JAMA 1987 Jul 3;258(1):67–74.

[9] Brixner D, Biltaji E, Bress A, Unni S, Ye X, Mamiya T, Ashcraft & J K. Journal of Medical Economics: The effect of pharmacogenetic profiling with a clinical decision support tool on healthcare resource utilization and estimated costs in the elderly exposed to polypharmacy. Biskupiak (2016) 19:3, 213–228.

[10] Rodolphe Meyer. Une approche économétrique pour l’analyse de l’impact médico-économique des systèmes d’information hospitaliers. Santé publique et épidémiologie. Université Pierre et Marie Curie - Paris VI, 2010. Français.

[11] Benmimoune L. Une approche pour la conception de systèmes d’aide à la décision médicale basés sur un raisonnement mixte à base de connaissance. Université de Technologie de Belfort-Montbeliard; 2016.

[12] Moher D, Liberati A, Tetzlaff J, Altman DG. Preferred Reporting Items for Systematic Reviews and MetaAnalyses: The PRISMA Statement. PLoS Med 2009.

[13] Kaplan JG. The net present value of investments in health. Med Interface. 1996; 9(11): 94–6

[14] Stroetmann KA, Jones T, Dobrev A, et al. eHealth is worth it: the economic benefits of implemented eHealth solutions at ten European sites. Luxembourg: Office for Official Publication of the European Communities, 2006: 13–30.

[15] Kaushal R, Jha AK, Franz C, Glaser J, Shetty KD. Return on investment for a computerized physician order entry system. J Am Med Inform Assoc. 2006; 13(3): 365–7.

[16] Loriane F. Le retour sur investissement du déploiement du Dossier Patient Informatisé: l’exemple du CHU d’Angers. 2009;103.

[17] Wu RC, Laporte A, Ungar WJ. Cost-effectiveness of an electronic medication ordering and administration system in reducing adverse drug events. J Eval Clin Pract 2007;13:440e8.

[18] Karnon J, McIntosh A, Dean J, et al. Modelling the expected net benefits of interventions to reduce the burden of medication errors. J Health Serv Res Policy 2008;13:85e91.

[19] Rosser WW, Hutchison BG, McDowell I, et al. Use of reminders to increase compliance with tetanus booster vaccination. CMAJ 1992;146:911e17. 10

[20] Fretheim A, Aaserud M, Oxman AD. Rational prescribing in primary care (RaPP): economic evaluation of an intervention to improve professional practice. PLoS Med 2006;3:e216.

[21] Plaza V, Cobos A, Ignacio-Garcia JM, et al. [Cost-effectiveness of an intervention based on the Global [Nitiative for Asthma (GINA) recommendations using a computerized clinical decision support system: a physicians randomized trial]. Med Clin (Barc) 2005;124:201e6.

[22] Chisolm DJ, McAlearney AS, Veneris S, et al. The role of computerized order sets in pediatric inpatient asthma treatment. Pediatr Allergy Immunol 2006;17:199e206.

[23] Mekhjian HS, Kumar RR, Kuehn L, et al. Immediate benefits realized following implementation of physician order entry at an academic medical center. J Am Med Inform Assoc 2002;9:529e39

[24] Stone WM, Smith BE, Shaft JD, et al. Impact of a computerized physician orderentry system. J Am Coll Surg 2009;208:960e7

[25] Tierney WM, Miller ME, Overhage JM, et al. Physician inpatient order writing on microcomputer workstations. Effects on resource utilization. JAMA 1993;269:379e83.

[26] Meyer R. Une approche économétrique pour l’analyse de l’impact médico-économique des systèmes d’information hospitaliers. Université Pierre et Marie Curie - Paris VI; 2010.

[27] Beard N, Elo K, Hitt LM, Housman MG, Mansfield G. Information technology and hospital performance: An econometric analysis ofcosts and quality. PricewaterhouseCoopers 2007.

[28] Menon N, Lee B, Eldenburg L. Productivity of information systems in the healthcare industry, Information Systems Research. 2000; 11(1): 83–92

[29] Meyer R, Degoulet P, Omnes L. Impact of Health Care Information Technology on Hospital Productivity Growth: a Survey in 17 Acute University Hospitals. Medinfo 2007; 12(1): 203–7

[30] Osei-Brison KM, Ko M. Exploring the relationship between information technology investments and firm performance using regression splines analysis. Information & management. 2004; 42:1 –13

